# Symptom Management When Non-Invasive Advanced Respiratory Support is Used During End-of-Life Care: A Systematic Review

**DOI:** 10.1101/2022.03.29.22273098

**Authors:** David Wenzel, Lucy Bleazard, Coral Pepper, Christina Faull

## Abstract

**Objectives:** to narrate the canon of knowledge around symptom control at end of life for patients using, or having recently used, non-invasive advanced respiratory support (NARS) at end of life for respiratory failure.

**Methods:** A systematic review forming a narrative synthesis from a wide range of sample papers from Medline, Embase, CINAHL, Emcare, Cochrane and OpenGrey databases. A secondary search of grey literature was also performed with hand searching reference lists and author citations. The review was undertaken using the ENTREQ checklist for quality.

**Results:** In total 15 studies were included in the synthesis and four themes were generated: NARS as a buoy (NARS can represent hope and relief from the symptoms of respiratory failure), NARS as an anchor (NARS brings significant treatment burden), Impact on Staff (uncertainty over the balance of benefit and burden as well as complex patient care drives distress amongst staff providing care) and the Process of Withdrawal (withdrawal of therapy felt to be futile exists as discrete event in patient care but is otherwise poorly defined).

**Conclusion:** NARS represents a complex interplay of hope, symptom control, unnaturally prolonged death and treatment burden. The literature captures the breadth of these issues but further, detailed, research is required in almost every aspect of practice around end-of-life care and NARS – especially how to manage symptoms at the end of life.

## INTRODUCTION

The use of non-invasive advanced respiratory support (NARS) is an established component of clinical practice and has become more prevalent over the last 20 years both in the acute hospital and community/home settings[1]. Non-invasive advanced respiratory support may be given via a facial interface in the form of continuous positive pressure (CPAP) or bilevel pressure (non-invasive ventilation/NIV/Bipap) and now has an evidence-based role in heart failure, obesity hypoventilation syndrome, sleep apnoea, neuromuscular disorders (including motor neurone disease) and COPD[2]. More recently it has evolved to support patients with COPD who have persistent hypercapnia, reducing the need for admission to hospital and increasing survival[3]. During the COVID-19 pandemic NARS was used extensively to treat respiratory failure due to COVID-19 related airways disease.

For many patients in respiratory failure NARS provides a successful outcome, that is that they survive the respiratory failure that would otherwise have caused their death, but despite intervention with NARS mortality rates remain high. In Covid-19 related disease, progression to CPAP is associated with an inpatient mortality rate of 85% amongst the over 65’s[4]. Patients with COPD have a 12% 90 day mortality rate following an admission, this mortality is two and a half times higher if they needed to receive NIV[5]. The effective management of these patients who deteriorate and die despite NARS thus requires consideration.

The British Thoracic Society’s (BTS) clinical standards report acknowledges mask removal (withdrawal of treatment) may be preferable but good quality palliation can be achieved with the interface in place[2]. Qualitative research exploring treatment burden and symptom relief with patients who received NIV for COPD and survived indicated that the mask may be uncomfortable[6] but it does bring significant relief from breathlessness[7]. There is, however, a lack of detailed, evidence-based guidance for practice. Exploration of the literature is needed that considers the care of patients who continue with NARS until death and where NARS is stopped/withdrawn to understand what symptoms occur, how can they be effectively managed, how long until death can be expected and how should patients be supported throughout this process.

As there are complex ethical considerations around withdrawing therapy[8] the preparation and impact on staff also requires consideration and how best to support staff during this process is ill-defined.

This review aims to narrate the canon of knowledge around symptom control at end of life for patients using, or having recently used, NARS at end of life for respiratory failure. A secondary outcome of understanding the impact this care has on the staff who provide it will also be reviewed. The review was undertaken using the ENTREQ checklist for quality[9].

## METHODS

The search included electronic databases as well as manually searching reference lists and citations. Medline, Embase, CINAHL, Emcare, Cochrane and OpenGrey databases were searched, and an grey literature search was also performed by reviewing relevant articles reference lists and citation searching. The Medline search strategy, which was adapted for other databases is included in appendix A.

Search terms followed the PCO model – population (adult patients, aged 18+, patients with respiratory failure), context (receiving non-invasive advanced respiratory support (CPAP, BiPAP, NIV), critically ill patients or those who did not survive the study), outcome (end of life care, symptom control).

Inclusion Criteria: qualitative, mixed methods or quantitative, examining symptom control when dying or critically ill, using or having recently used NARS.

Exclusion Criteria: paediatric patients, invasive ventilation with sedation (i.e. intubated in intensive care) or included patients using only oxygen (including high flow).

### ARTICLE IDENTIFICATION

2987 articles were identified through the initial search with a further 3 articles identified from reference lists. After deduplication 2195 records remained (reviewed by DW), 2076 of which did not meet the inclusion criteria or met the exclusion criteria. 119 full texts were assessed for eligibility by DW and LB, 100 of which were excluded for inadequate focus on symptom control needs. 4 articles were identified as likely suitable but full texts could not be obtained. In each instance the correspondence authors were contacted on their given emails. Three did not reply and one was unable to pass on the full text. 15 studies were included in the review (figure 1) and are summarised in table 1.

Given the heterogeneity of data included the studies were appraised against the inclusion criteria. The rigor of qualitative studies was appraised using the critical appraisals skills programme (CASP) checklist[10].

**Figure 1.**
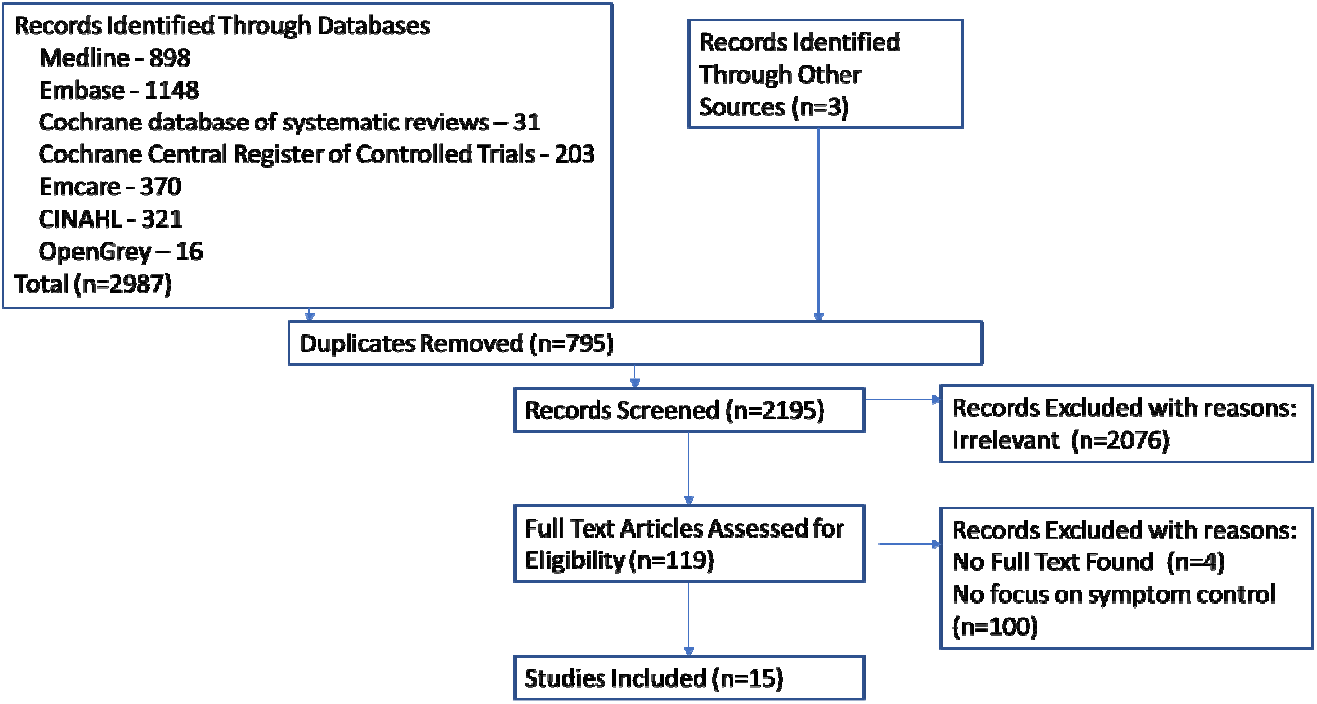

### DATA EXTRACTION AND SYNTHESIS

An approach of narrative synthesis was used[11] to understand the journey of critically ill patients using NARS who did not survive their admissions. A grounded theory approach was then adopted to establish a meta thematic analysis of themes that arched over the included papers. Papers were uploaded into a qualitative data management system (MAXQDA Plus 2020 20.4.0). Data under the headings, ‘results’, ‘discussion’ and ‘conclusion’ were included for coding. Coding was inductive and descriptive with generation of codes from the data itself. Papers were sequentially coded with headings from earlier papers and new codes generated as they occurred. When coding of all papers had been undertaken an iterative approach was used to apply later developed codes to earlier papers. Coding was primarily undertaken by DW. Thematic analysis was similarly inductive with a mixture of descriptive thematic generation and in vivo theme production. This was completed by DW and LB, with additional independent commentary on emerging codes and themes from CF.

**Table 1.**
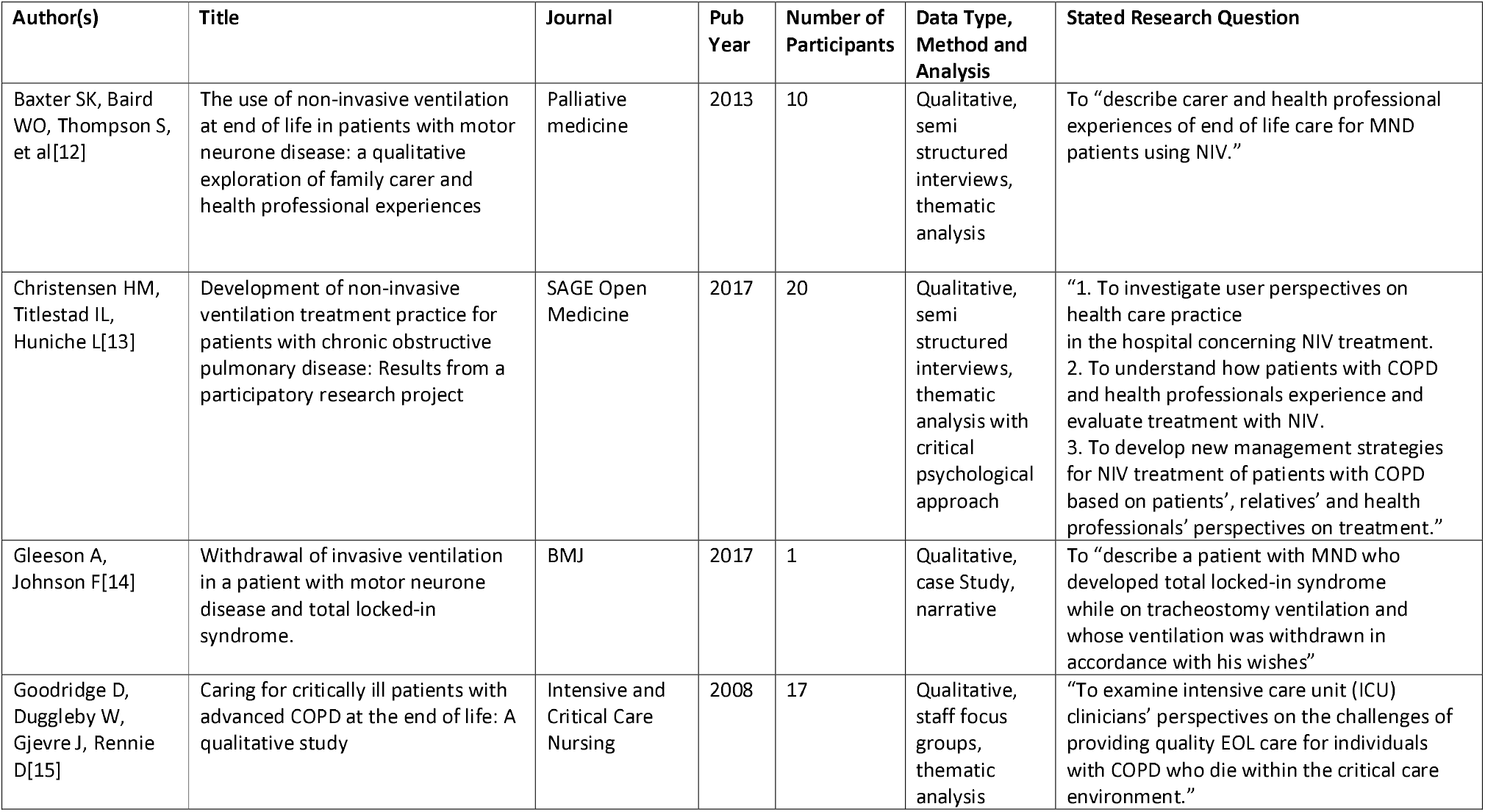
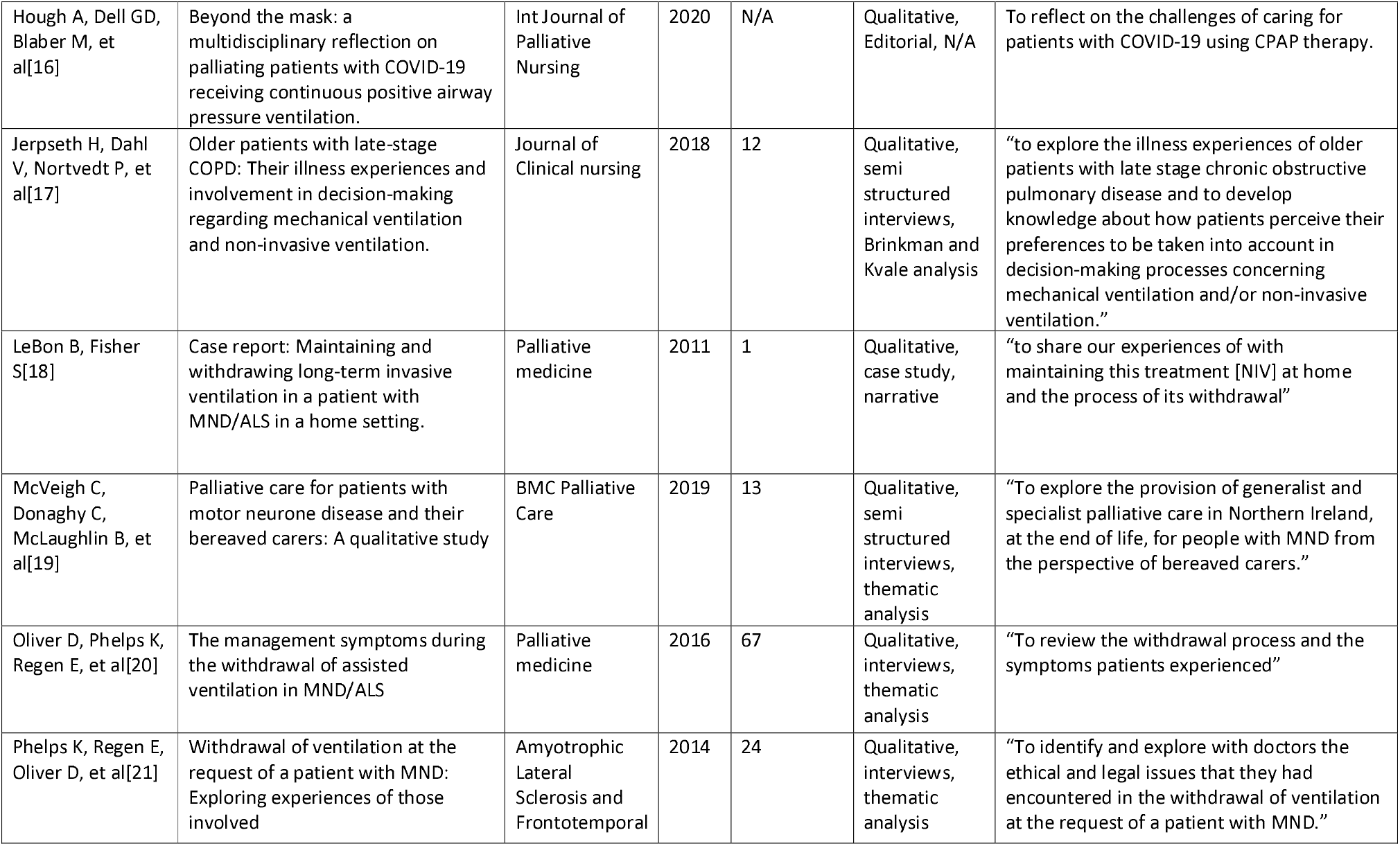
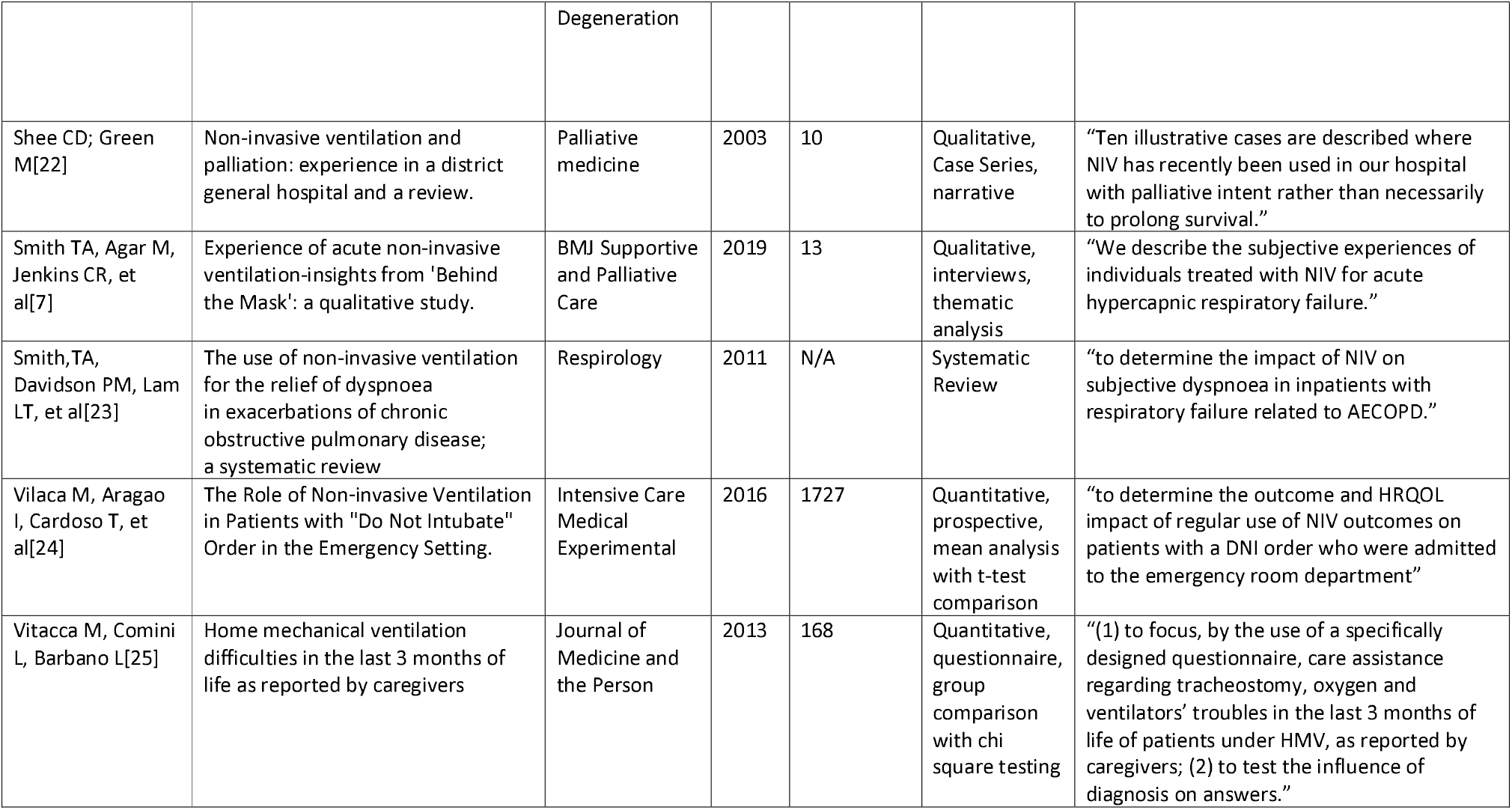

## FINDINGS

Of the 15 full text articles included in the data synthesis 12 were purely qualitative, two were quantitative and one review article was included. Papers spanned from 2003 to 2020 and utilised techniques including semi-structured interviews, case series, editorials and observational studies.

Four themes were developed from the data. The first two themes developed narrate the patient’s complex journey and relationship with NARS, ‘NARS as a buoy’, ‘NARS as an Anchor’. The third theme explored the impact providing care had on healthcare workers, ‘Impact on Staff’. Withdrawal as a discrete event within patient care was noted in several papers and formed its own theme, ‘The Process of Withdrawal’.

### NARS as a Buoy

Throughout the literature reviewed the primary focus for the initiation and use of NARS was for the preservation of life, conceptually referred to as life buoy in one paper. There was only limited consideration of NARS as a source of symptom relief but despite this many patients appeared to receive significant improvement in symptoms on initiation of NARS. This included improved breathlessness[7,12,22], anxiety[7,12,13,19] and insomnia. Resolution of insomnia was noted in two papers and often followed long periods of sleeplessness[7,22].

> *“Sleep deprivation prior to hospitalisation was frequently mentioned and participants expressed relief at being able to sleep once NIV started”[7]*

NARS also represented a symbol of hope to many patients[7,13,17]. An active treatment that may buy time and another chance of life[12,22]. This was described in one paper, conceptually, as a buoy:

> *“They expressed an appreciation for having the opportunity to use the mask as a “life buoy”—a symbol of hope and survival —even when there was no prospect of healing*.*”[17]*

Two papers, that focused on patients who had survived episodes where NIV had been used in the context of an acute hospital admission for respiratory failure, reported a general acceptance of future NIV use[7,13]. Demonstrating that the use of NIV is viewed positively, albeit in the context of having survived. Accurately assessing treatment burden may be complicated by viewing the acute episodes in the context of having survived (with a preserved quality of life[24]). Retrospective analysis of these episodes may be further complicated by poor recollection of the acute episode[13,7,17].

> *“It appeared in the interviews that 15 out of 16 patients and 4 relatives all expected and wanted to be treated with NIV despite displays of resistance”[13]*

### NARS as an Anchor

While there were potential symptom benefits to NARS these were counterbalanced by significant treatment burdens. NARS may represent an anchor which adds burden to the patient and which of these opposing pulls is stronger, the hope and relief of the buoy or the symptom burden of the anchor, is difficult to predict[16,19,22,24].

NARS was noted at times to contribute the breathlessness it was intended to treat[7,13,17,19], though this may improve with adjustment of settings and perseverance beyond initiation[7,19].

> *“Feeling strangled by the NIV mask caused anxiety. Not being able to think straight, made them focus on breathing as a main issue*.*”[13]*

Patients experiencing a critical respiratory illness are prone to anxiety[15,16,19,22,25] but NARS can further exacerbate this[7,17,19,22]. This may be driven by worsening breathlessness but is also contributed to by feelings of claustrophobia related to the interface[13,16,17,22] and the noise of the machine[15,16,17,22]. The noise of NARS and the mask created issues in communication which complicated all aspects of care.

> *“At times, it felt dehumanising to have to almost shout at patients to be heard”[16]*

The mask impacted the ability of healthcare workers to provide basic nursing care, such as mouthcare[12,16].

> *“She got so distressed without the mask support that her mouth was in a terrible state. You couldn’t get in to give her proper mouth care because it meant taking the mask off for periods of time”[12]*

The nature of the mask interface was noted commonly to cause symptom burden[7,12,13,16,17,22].

### Impact on Staff

Patients in receipt of NARs had a significant risk of mortality with high symptom burdens[24]. The nature of this work, combined with the uncertain beneficence[19,22,24] and potential maleficence of NARS[7,15,16,17,19,22] negatively impacted staff[15,16,18,19,21].

> *“And that’s where it becomes hard on our part, because you know, pretty well, what’s coming, and we’re just simply prolonging[death], and at times feel that you’re being abusive*.*”[15]*

Almost half of the papers referenced the impact that providing care had on healthcare professionals and much of this was related to uncertainty. The difficulties with prognosticating, whether or not to remove the mask and the difficulty of controlling symptoms all featured in this professional unease which was compounded by a lack of good quality research[7,12,13,14,15,16,19,20,21,22,24].

> *“These participants described some uncertainty regarding how best to manage NIV in the final stage and whether usage should be withdrawn”[12]*
>
> *“[when] they’re… on bipap, and that again is an incredible struggle. And at that point, when you’re administering sedation pain medication, there’s that area, how much do you give?”[15]*

### The Process of Withdrawal

The decision to withdraw therapy was often not well explored in papers focusing on acute admissions, but was noted on several occasions to be removed due to patient preference[16,22]. Two papers regarded an increased dependence on ventilation for symptom control to indicate a worsening prognosis[12,16]. One paper described indications to withdraw which included continued deterioration and poor mask tolerance[16]. The process of withdrawal was poorly described but 6 papers referenced the use of anticipatory medications to control symptoms[12,14,16,18,20,22] and one paper acknowledged the need to replace the mask during withdrawal to allow for better symptom control[20].

> *“The implications of stopping treatment were explained to him. He was reviewed by the hospital palliative care nurse and started on a low-dose sc diamorphine infusion for breathlessness. When he felt comfortable he removed his NIV facemask, and he died peacefully on the ward later that night with his family present*.*”[22]*

Breathlessness and anxiety were noted as the primary symptoms of concern during mask withdrawal[20].

## DISCUSSION

This literature review aimed to explore symptom control at the end of life in the context of NARS use. 15 papers were included in the final analysis which contributed to four themes. The paucity of studies in this specific area makes drawing robust conclusions challenging. However, a narrative of uncertainty is present throughout what data there is.

NARS can be an agent of symptom control yet was rarely reported to be used in this way. Some patients, specifically those that survived, reported improved breathlessness, anxiety and sleep whilst using NARS during episodes of acute respiratory deterioration. The reported willingness to have NARS again following acute exacerbations may suggest a net benefit to the intervention. However, there is no clear answer as to whether this symptom control can be better achieved using less burdensome interventions.

There may exist a transition point at which NARS changes from being a buoy to an anchor. If this point is not recognised this may contribute to the feelings of futility that healthcare workers reported following the death of patients on NARS – if the interface continued to provide good symptom control it is unlikely this feeling would exist.

In Vilaca et al[24] a cohort of 29 patients are commenced on NIV for symptom control intent only. This group, who almost all died on the first day of admission, had an almost 60% discontinuation rate – possibly suggesting that NARS is not effective enough symptom relief to offset its treatment burden. It is unclear how clinicians in the determined who had the poorest chance of survival and this may have influenced the study outcome with regards to drop-out from NARS use.

It is likely, supported by Smith 2019[7], that the balance of anchor and buoy shifts during the illness episode. Breathlessness induced by NARS seems to settle after a period of accommodation but increasing symptom burden can drive withdrawal decisions. It is unclear if anxiolytic medication may have a role to play in lessening the duration of the accommodation period. It is likely that constant reassessment of the opposing pulls of anchor and buoy is required, particularly as prognosis becomes more apparent.

A conspicuous absence in the literature was information about the process of withdrawal of NARS. Enough papers referenced the use of anticipatory medication to suggest that NARS withdrawal isn’t quite a simple as just taking the mask off. Yet there were no detailed accounts, other than in the context of elective withdrawal in patients with MND[20], about how NARS may be withdrawn with adequate symptom management. There were commonalities to the burden of NARS (claustrophobia, noise, difficulty in communication), but clear distinctions in the literature reviewed between the issues of electively withdrawing long term assisted ventilation in MND and deciding to withdraw ineffective or unwanted NARS in acute episodes of COPD or COVID – primarily that opportunity to plan and prepare are markedly different.

Such a wide area of uncertainty provides opportunities for further research. Before it is possible to intervene and improve the withdrawal process it is necessary to understand the processes, complexities and outcomes of common practice.

What is understood in the current literature is that this practice has an impact on the staff involved in the process[15]. Much of this impact could be viewed through the lens of moral injury[26] whereby healthcare workers become witnesses to acts that affront their conscience. Often by being forced to commit an act they do not believe in, such as providing high burden NARS treatment in the context of a limited prognosis[15]. This area of research has become more pertinent since the advent of the pandemic, where NARS withdrawals moved from being infrequent winter respiratory care events to daily general medical ward events[27]. Further research on how to mitigate such impacts and how to support, staff would be valuable in safeguarding our healthcare workers.

It is unclear from the available data how the symptom burden differs between patients with hypercapnic and hypoxic respiratory failure and this may warrant further exploration. This review also noted a lack research related to of family or lay caregivers who may be able to act as more accurate observers of the end-of-life period. This too may provide a further avenue to explore the experiences and needs of those dying using, or having recently used, NARS.

## CONCLUSION

NARS represents a complex interplay of hope, symptom control, unnaturally prolonged death and treatment burden. The literature captures the breadth of these issues but further, detailed, research is required in almost every aspect of practice around end-of-life care and NARS – especially how to manage symptoms at the end of life. What is clear is that staff are witnesses to this difficult and uncertain time. The impact it has on them is poorly defined but supporting them is likely to be essential if the frequency of NARS interventions increases.

## Supporting information

ENTREQ checklist

## Data Availability

All data produced in the present study are available upon reasonable request to the authors

## Appendix A

### Medline strategy – Experiences of dying using NIV

**Run 17**^th^ **February 2021**

**898 results**

1. Lung Diseases, Obstructive/
2. exp Pulmonary Disease, Chronic Obstructive/
3. emphysema*.tw.
4. (chronic* adj3 bronchiti*).tw.
5. (obstruct* adj3 (pulmonary or lung* or airway* or airflow* or bronch* or respirat*)).tw.
6. COPD.tw.
7. exp Respiratory Insufficiency/
8. (respiratory adj3 (fail* or insufficien*)).tw.
9. Hypoxia/
10. hypoxi*.tw.
11. hypercapni*.tw.
12. Hypercapnia/
13. (interstitial adj2 (“lung disease*” or pneumoni*)).tw.
14. exp Lung Diseases, Interstitial/
15. ILD.tw.
16. exp Coronavirus/
17. Coronavirus Infections/
18. ((corona* or corono*) adj1 (virus* or viral* or virinae*)).tw.
19. (coronavirus* or coronovirus* or coron?virinae* or “2019-nCoV” or 2019nCoV or “2019-CoV” or nCoV2019 or “nCoV-2019” or “COVID-19” or COVID19 or “CORVID-19” or CORVID19 or “WN-CoV” or WNCoV or “HCoV-19” or HCoV19 or CoV or “2019 novel*” or Ncov or “n-cov” or “SARS-CoV-2” or “SARSCoV-2” or “SARSCoV2” or “SARS-CoV2” or SARSCov19 or “SARS-Cov19” or “SARSCov-19” or “SARS-Cov-19” or “SARSr-cov” or Ncovor or Ncorona* or Ncorono* or NcovWuhan* or NcovHubei* or NcovChina* or NcovChinese* or Wuhan virus* or novel CoV or CoV 2 or CoV2 or betacoron?vir*).tw.
20. ((heart or cardiac or myocard*) adj2 (fail* or insufficien* or decomp*)).tw.
21. exp Heart Failure/
22. (pulmonary adj (oedem* or edem*)).tw.
23. (cardiogenic adj (oedem* or edem*)).tw.
24. Pulmonary Edema/
25. bronchiect*.tw.
26. exp Bronchiectasis/
27. (neuromuscular adj2 (disease* or disorder*)).tw.
28. exp Neuromuscular Diseases/
29. (“motor neuron*” adj disease*).tw.
30. MND.tw.
31. amyotrophic lateral sclerosis.tw.
32. “progressive bulbar pals*”.tw.
33. pseudobulbar pals*.tw.
34. progressive muscular atroph*.tw.
35. primary lateral sclerosis.tw.
36. monomelic amyotrophy.tw.
37. 1 or 2 or 3 or 4 or 5 or 6 or 7 or 8 or 9 or 10 or 11 or 12 or 13 or 14 or 15 or 16 or 17 or 18 or 19 or 20 or 21 or 22 or 23 or 24 or 25 or 26 or 27 or 28 or 29 or 30 or 31 or 32 or 33 or 34 or 35 or 36
38. death*.ti,kw.
39. die.tw.
40. dying.tw.
41. passed away.tw.
42. deceased.tw.
43. “end of life”.tw,kw.
44. palliat*.tw,kw.
45. terminal*.tw.
46. hospice*.tw,kw.
47. exp Terminal Care/
48. “Hospice and Palliative Care Nursing”/
49. Palliative Care/
50. Palliative Medicine/
51. Death/
52. 38 or 39 or 40 or 41 or 42 or 43 or 44 or 45 or 46 or 47 or 48 or 49 or 50 or 51
53. “non-invasive ventilation”.tw,kw.
54. noninvasive ventilation.tw,kw.
55. pulmonary ventilation.tw.
56. positive pressure ventilation.tw.
57. pressure support ventilation.tw.
58. positive end expiratory pressure*.tw.
59. NIV.tw,kw.
60. NIAV.tw,kw.
61. NIPPV.tw,kw.
62. NPPV.tw,kw.
63. mechanical ventilation.tw.
64. artificial ventilation.tw.
65. assisted ventilation.tw.
66. artificial respiration.tw.
67. continuous positive airway pressure.tw,kw.
68. CPAP.tw,kw.
69. nCPAP.tw,kw.
70. bilevel positive airway pressure.tw,kw.
71. “bi-level positive airway pressure”.tw,kw.
72. biphasic positive airway pressure.tw,kw.
73. “bi-phasic positive airway pressure”.tw,kw.
74. BiPAP.tw,kw.
75. BPAP.tw,kw.
76. Noninvasive Ventilation/
77. exp Positive-Pressure Respiration/
78. Respiration, Artificial/
79. 53 or 54 or 55 or 56 or 57 or 58 or 59 or 60 or 61 or 62 or 63 or 64 or 65 or 66 or 67 or 68 or 69 or 70 or 71 or 72 or 73 or 74 or 75 or 76 or 77 or 78
80. (view* or perspective* or thought* or feel* or experience* or reflect* or attitude* or receptive* or response* or satisf*9 or benefit* or preferenc*).tw,kw.
81. support*.tw,kw.
82. symptom*.tw,kw.
83. palliat*.tw,kw.
84. ((patient* or person* or carer* or famil*) adj2 need*).tw.
85. qualitative*.tw.
86. Patient Satisfaction/
87. exp Qualitative Research/
88. 80 or 81 or 82 or 83 or 84 or 85 or 86 or 87
89. 37 and 52 and 79 and 88

