## Supplementary material for "Symptom Management When Non-Invasive Advanced Respiratory Support is Used During End-of-Life Care: A Systematic Review": ENTREQ checklist

| **The ENTREQ Checklist Item** | **Guide and description** | **Reported on page #** |
| --- | --- | --- |
| Aim | State the research question the synthesis addresses | 2 |
| Synthesis methodology | Identify the synthesis methodology or theoretical framework which underpins the synthesis, and describe the rationale for choice of methodology (e.g. meta-ethnography, thematic synthesis, critical interpretive synthesis, grounded theory synthesis, realist synthesis, meta-aggregation, meta-study, framework synthesis). | 3 |
| Approach to searching | Indicate whether the search was pre-planned (comprehensive search strategies to seek all available studies) or iterative (to seek all available concepts until theoretical saturation is achieved). | 2 |
| Inclusion criteria | Specify the inclusion/exclusion criteria (e.g. in terms of population, language, year limits, type of publication, study type). | 3 |
| Data sources | Describe the information sources used (e.g. electronic databases (MEDLINE, EMBASE, CINAHL, psychINFO, Econlit), grey literature databases (digital thesis, policy reports), relevant organisational websites, experts, information specialists, generic web searches (Google Scholar), hand searching, reference lists) and when the searches were conducted; provide the rationale for using the data sources. | 2 |
| Electronic Search strategy | Describe the literature search (e.g. provide electronic search strategies with population terms, clinical or health topic terms, experiential or social phenomena related terms, filters for qualitative research and search limits). | 2, 3 and Appendix A |
| Study screening methods | Describe the process of study screening and sifting (e.g. title, abstract and full text review, number of independent reviewers who screened studies) | 3 |
| Study characteristics | Present the characteristics of the included studies (e.g. year of publication, country, population, number of participants, data collection, methodology, analysis, research questions). | 4,5,6 |
| Study selection results | Identify the number of studies screened and provide reasons for study exclusion (e.g. for comprehensive searching, provide numbers of studies screened and reasons for exclusion indicated in a figure/flowchart; for iterative searching describe reasons for study exclusion and inclusion based on modifications t the research question and/or contribution to theory development). | 3 |
| Rationale for appraisal | Describe the rationale and approach used to appraise the included studies or selected findings (e.g. assessment of conduct (validity and robustness), assessment of reporting (transparency), assessment of content and utility of the findings). | 3 |
| Appraisal items | State the tools, frameworks and criteria used to appraise the studies or selected findings (e.g. Existing tools: CASP, QARI, COREQ, Mays and Pope [25]; reviewer developed tools; describe the domains assessed: research team, study design, data analysis and interpretations, reporting). | 3 |
| Appraisal process | Indicate whether the appraisal was conducted independently by more than one reviewer and if consensus was required. | 3 |
| Appraisal results | Present results of the quality assessment and indicate which articles, if any, were weighted/excluded based on the assessment and give the rationale. | 3 |
| Data extraction | Indicate which sections of the primary studies were analysed and how were the data extracted from the primary studies? (e.g. all text under the headings “results /conclusions” were extracted electronically and entered into a computer software). | 3 |
| Software | State the computer software used, if any. | 3 |
| Number of reviewers | Identify who was involved in coding and analysis. | 3 |
| Coding | Describe the process for coding of data (e.g. line by line coding to search for concepts). | 3 |
| Study comparison | Describe how were comparisons made within and across studies (e.g. subsequent studies were coded into pre-existing concepts, and new concepts were created when deemed necessary). | 3 |
| Derivation of themes | Explain whether the process of deriving the themes or constructs was inductive or deductive. | 3 |
| Quotations | Provide quotations from the primary studies to illustrate themes/constructs, and identify whether the quotations were participant quotations or the author’s interpretation | 7,8 |
| Synthesis output | Present rich, compelling and useful results that go beyond a summary of the primary studies (e.g. new interpretation, models of evidence, conceptual models, analytical framework, development of a new theory or construct). | 8,9 |
